# Epigenetic clocks and longitudinal plasma biomarkers of Alzheimer’s disease

**DOI:** 10.1101/2025.08.12.25333453

**Authors:** Bowei Zhang, Linda K. McEvoy, Steve Nguyen, Mark A. Espeland, Stephen R. Rapp, Steve Horvath, Ake Lu, Andrea Z. LaCroix, Caroline M. Nievergelt, Adam X. Maihofer, Susan M. Resnick, Michelle M. Mielke, Kenneth Beckman, Danni Li, Brian Silver, JoAnn E. Manson, Luigi Ferrucci, Aladdin H. Shadyab

## Abstract

**INTRODUCTION:** Chronological age is the strongest risk factor for Alzheimer’s disease and related dementias (ADRD). However, the association of accelerated biological aging relative to chronological age with ADRD pathology is unclear.

**METHODS:** In a cohort of 2,366 cognitively unimpaired older women from the Women’s Health Initiative Memory Study, we examined associations of five baseline measures of epigenetic age acceleration (EAA) with 15-year changes in plasma ADRD biomarkers.

**RESULTS:** At baseline, higher AgeAccelPheno was associated with lower amyloid-β42 to amyloid-β40 (Aβ42:Aβ40) ratio, and higher AgeAccelGrim2 was associated with elevated neurofilament light (NfL). Longitudinally, higher DunedinPACE – which measures the pace of biological aging – was associated with faster increases in phosphorylated tau at threonine 181 (p-tau181), p-tau217, NfL, and glial fibrillary acidic protein (GFAP) over 15 years.

**DISCUSSION:** Accelerated biological aging, particularly as indicated by DunedinPACE, was associated with increasing levels of plasma ADRD biomarkers over time.

## 1 BACKGROUND

Although chronological age is the strongest risk factor for Alzheimer’s disease and related dementias (ADRD), it remains unclear whether accelerated biological aging relative to chronological age is associated with ADRD pathology.^1^ Epigenetic clocks are DNA methylation (DNAm)-based biomarkers measuring an individual’s biological age.^2^ Several distinct epigenetic clocks have been developed and validated, including first-generation clocks trained to predict chronological age, such as those developed by Horvath and Hannum, second-generation clocks developed to predict clinical phenotypes (PhenoAge) and mortality (GrimAge/GrimAge2), and a third-generation clock developed to predict the pace of biological aging across different organ systems (DunedinPACE).^3–8^ Epigenetic age acceleration (EAA), indicating faster biological aging relative to chronological age, has been associated with higher risk of age-related diseases and all-cause mortality.^2–9^ Longitudinal studies in multiple cohorts, including the Women’s Health Initiative Memory Study (WHIMS), have shown that EAA is associated with greater cognitive decline.^10–15^ EAA has also been associated with increased risk of mild cognitive impairment (MCI) and dementia in some studies.^16–19^

Over the past decade, significant progress has been made in identifying blood-based biomarkers of ADRD that are non-invasive and more cost-effective compared to cerebrospinal fluid and positron emission tomography in vivo measures of ADRD pathology.^20–26^ These biomarkers include plasma biomarkers of AD-specific pathology, including amyloid-β 42 (Aβ42), amyloid-β 40 (Aβ40), and phosphorylated tau at threonine 181 (p-tau181) and at threonine 217 (p-tau217). Non-specific markers of neuronal injury (neurofilament light chain protein [NfL]) and of neuroinflammation (glial fibrillary acidic protein [GFAP]) have also been associated with increased dementia risk.^22,27–29^ Only one prior study has examined associations of epigenetic clocks of biological aging with plasma biomarkers of ADRD.^30^ However, that study was limited by its cross-sectional design; examined only Hispanic/Latino adults, thereby limiting generalizability to other racial and ethnic groups; did not examine changes in plasma biomarkers over time; and did not evaluate plasma p-tau217. A better understanding of whether accelerated biological aging is associated with elevated levels of plasma biomarkers of ADRD over time could help identify high-risk populations and inform precision medicine approaches targeting specific aging pathways to prevent ADRD.

In this study, we examined associations of baseline EAA measured using several well-studied epigenetic clocks with plasma ADRD biomarkers including Aβ42:Aβ40 ratio, p-tau181, p-tau217, NfL, and GFAP. Importantly, we had plasma biomarker data available from two visits an average of 15 years apart, allowing us to examine the extent to which EAA predicts longitudinal changes in plasma ADRD biomarkers. We examined several different clocks that capture different aspects of biological aging. We hypothesized that EAA would be associated with greater indication of ADRD pathology at baseline and greater changes in markers of pathology over time (i.e., lower Aβ42:Aβ40 and higher p-tau181, p-tau217, NfL, and GFAP). We hypothesized that the second– and third-generation clocks would show greater association with EAA than the first-generation clocks and that the strongest associations would be found for DunedinPACE, as we previously found that this epigenetic clock showed the strongest association with cognitive decline in the WHIMS cohort.^10^

## 2 METHODS

### 2.1 Study Design and Population

WHIMS was an ancillary study of the Women’s Health Initiative hormone therapy trials. WHIMS was designed to investigate the effects of hormone therapy on cognitive outcomes among 7,479 women ages 65-79 years at enrollment who were cognitively unimpaired at randomization in 1996-1999. Details on the WHIMS design and protocols have been published.^31–33^ Women were randomized either to conjugated equine estrogens (CEE) plus medroxyprogesterone acetate (MPA) vs placebo among women with an intact uterus and CEE alone vs placebo among women with prior hysterectomy. The trials were stopped in 2002 and 2004, respectively, but follow-up for outcomes continued. Annual follow-up for in-person cognitive assessments continued through 2007. In 2008, WHIMS transitioned to annual telephone-administered cognitive assessments in the WHIMS Epidemiology of Cognitive Health Outcomes (WHIMS-ECHO) study, which followed participants for cognitive outcomes through 2021.^34^

Among 7,479 WHIMS participants, we excluded 240 with only one WHIMS cognitive assessment, 519 who did not consent to DNA sharing, and 304 with no baseline DNA or buffy coat available, leaving 6,416 participants whose DNA was extracted from blood samples collected at the baseline visit (1996-1999) and shipped to the University of Minnesota Genomics Center for DNA methylation measurement. DNAm was measured using the Illumina Infinium MethylationEPIC v2.0 using standard protocols. After quality control (see Supplementary Material), baseline epigenetic data were available for 6,069 participants. For the present analysis, we further selected women with data on plasma biomarkers of ADRD available at baseline for cross-sectional analyses (N = 2,366) or both baseline and a second time point an average of 15 years later for longitudinal analyses (N = 873). This study was approved by the Institutional Review Board at University of California San Diego. All participants provided written informed consent.

### 2.2 Plasma Biomarkers

Fasting blood was drawn at baseline and a second time point an average of 15 years later. Samples were processed, frozen at –70°C, and then shipped to a repository in Rockville, MD maintained by Fisher Bioservices. All plasma biomarker assays were performed at the Advanced Research and Diagnostics Laboratory (ARDL) at the University of Minnesota in 2024. Samples were shipped to the laboratory on dry ice. Levels of plasma biomarkers were measured using the Quanterix HD-X platform. The Simoa® Human Neurology 4-Plex E platform was used to measure levels of Aβ40, Aβ42, NfL, and GFAP. P-tau181 was measured using the Simoa® p-tau181 v2 assay. P-tau217 was measured using the ALZpath Simoa® pTau-217 v2 assay. Samples were assayed in singlets with the inclusion of 192 duplicates; laboratory personnel were blinded to the inclusion of these duplicate samples, and they were also blinded to cognitive impairment status. Baseline and follow-up samples were measured in the same laboratory at the same time using a single lot of reagents for each biomarker. The average intra-assay coefficients of variation for Aβ40, Aβ42, NfL, GFAP, p-tau181, and p-tau217 derived from these duplicates were 9.2%, 9.3%, 7.1%, 9.8%, 10.2%, and 11.4%, respectively.

The following inter-assay laboratory coefficients of variation (CVs) were derived from an Advanced Research and Diagnostics Laboratory pooled sample and the two kit controls, which were run on every plate along with the samples: 1) Aβ40: 4.8%, 5.7%, and 13.3% at mean concentrations of 19.0, 97.6, and 42.0 pg/mL, respectively; 2) Aβ42: 3.6%, 4.9%, and 12.8% at mean concentrations of 8.3, 38.0, and 2.4 pg/mL, respectively; 3) NfL: 6.4%, 9.0%, and 9.4% at mean concentrations of 23.5, 499.2, and 8.2 pg/mL, respectively; 4) GFAP: 10.6%, 9.8%, and 15.7% at mean concentrations of 188.4, 3744.0, and 72.3 pg/mL, respectively; 5) p-tau181: 7.2%, 6.4%, and 10.7% at mean concentrations of 42.4, 1130.9, and 14.9 pg/mL, respectively; and 6) p-tau217: 11.4%, 11.2%, and 12.9% at mean concentrations of 0.75, 0.39, and 0.15 pg/mL, respectively.

### 2.3 Epigenetic Clocks

We examined EAA using the first-, second-, and third-generation epigenetic clocks that are the most widely studied: AgeAccelHorvath, AgeAccelHannum, AgeAccelPheno, AgeAccelGrim2, and DunedinPACE. AgeAccelHorvath, AgeAccelHannum, AgeAccelPheno, and AgeAccelGrim2 were calculated with the online Horvath and Clock Foundation DNAm Age Calculator (https://dnamage.clockfoundation.org/). DunedinPACE was calculated using R code available at GitHub from the Belsky Lab (https://github.com/danbelsky/DunedinPACE).

AgeAccelHorvath, derived from the multi-tissue Horvath epigenetic clock, and AgeAccelHannum, derived from the blood-based Hannum epigenetic clock, are first-generation clocks developed on chronological age.^3,4^ AgeAccelPheno is a second-generation clock capturing phenotypic age, consisting of age and 9 clinical biomarkers (e.g., levels of albumin, creatinine, glucose, C-reactive protein, and immune markers); it has been shown to outperform the first-generation clocks in predicting healthspan indices.^5^ AgeAccelGrim2, also a second-generation clock, is a composite biomarker of DNAm-based surrogates of plasma proteins (e.g. cystatin C, CRP, GDF-15), a DNAm-based estimator of smoking pack years, age, and sex.^7^ DunedinPACE, Dunedin (P)ace of (A)ging (C)alculated from the (E)pigenome, a third-generation clock, was developed using longitudinal data from 19 biomarkers assessing cardiovascular, metabolic, renal, hepatic, immune, dental, and pulmonary systems to measure the pace of biological aging across organ systems among middle-aged adults followed for 20 years and has been shown to predict morbidity, disability, and mortality.^8^

### 2.4 Covariates

Baseline questionnaires assessed age, race, ethnicity, education, smoking status, diabetes (defined as ever having been diagnosed and treated with pills or insulin shots), cardiovascular disease (CVD) including myocardial infarction and stroke), and total energy expenditure from recreational physical activity (in metabolic equivalent [MET] hours/week). Hypertension was defined as either self-report of physician-diagnosed hypertension, use of hypertensive medications, or measured systolic blood pressure ≥130 mm Hg or diastolic blood pressure ≥80 mm Hg. Height and weight were measured with a stadiometer and balance beam scale, respectively, to calculate body mass index (BMI; kg/m^2^). Apolipoprotein E epsilon 4 (*APOE* ε4) carrier status, defined as presence of at least one ε4 allele versus non-carrier, was determined in women with available genome-wide genotyping data based on 2 single nucleotide variants, rs429358 and rs7412. Imputation was performed using the 1000 Genomics Project reference panel and the MaCH algorithm implemented in Minimac.^35^ Both single nucleotide polymorphisms (SNPs) had high imputation quality (R^2^ >0.97 for rs429358 and R^2^>0.97 for rs7412).^10^ Hormone therapy treatment arm in the original WHIMS trial (estrogen alone, estrogen placebo, estrogen plus progestin, or estrogen plus progestin placebo) was also included as a covariate. Fasting blood samples from previous WHI ancillary studies were sent to the University of Minnesota Medical Center Advanced Research and Diagnostics Lab for measurement of serum lipids including low-density (LDL; mg/dL) and high-density (HDL; mg/dL) lipoprotein cholesterol. HDL was measured using the HDL-C plus third-generation direct method on the Roche Modular P Chemistry Analyzer. LDL was calculated using the Friedewald formula.^36^ Creatinine was assayed using the Creatinine Plus Reagent (Roche Diagnostics) on the Modular P Chemistry Analyzer (Roche Diagnostics). We calculated estimated glomerular filtration rate (eGFR) using the 2009 Chronic Kidney Disease Epidemiology Collaboration equation based on serum creatinine measurements.^37^ White blood cell (WBC) counts were estimated for CD4+T cells, CD8+ T cells, natural killer cells, B cells, monocytes, neutrophils, and granulocytes using IDOL for models with DunedinPACE or with the Houseman method for other epigenetic clocks.^38,39^

### 2.5 Statistical Methods

#### 2.5.1 Descriptive Statistics

We calculated the ratio of plasma Aβ42 to Aβ40, because the ratio is a better measure of amyloid pathology compared to either biomarker alone.^40^ Since plasma biomarkers were non-normally distributed, Aβ42:Aβ40, NfL, GFAP, p-tau181, and ptau-217 were standardized using log2-based Z-transformation in all analyses, consistent with prior literature.^41^

Means and standard deviations (SDs), or counts and proportions, were reported for continuous and categorical baseline covariates, respectively, across quartiles of DunedinPACE, which was the clock we were most interested in examining due its prior association with cognitive decline in WHIMS.^10^ Differences in continuous and categorical variables across DunedinPACE quartiles were examined using Kruskal-Wallis rank sum tests and Chi-square tests, respectively. Fisher’s exact tests with simulated p-values (based on 2,000 replicates) were used for categorical variables with low expected counts. Means and SDs of plasma biomarkers were reported for participants (N = 873) with longitudinal data.

#### 2.5.2 Associations of Epigenetic Age Acceleration with Plasma Biomarkers at Baseline

Associations of each epigenetic clock with plasma biomarkers at baseline were assessed by linear regression models to estimate β coefficients and their 95% confidence intervals for 1 SD increases in EAA. Minimally adjusted models controlled for the following potential confounders: chronological age, hormone therapy trial arm, education, smoking status, race, and ethnicity. Fully adjusted models additionally controlled for physical activity, BMI, diabetes, CVD, hypertension, total cholesterol, HDL cholesterol, eGFR, and estimated blood cell composition (CD8 T, CD4 T, natural killer, B cell, monocyte, neutrophil). These potential confounders were selected from prior literature and include factors that may influence levels of plasma ADRD biomarkers (eGFR, BMI, co-morbidities, cholesterol levels).^10,41^ Models also incorporated inverse propensity score weights to account for sample selection for plasma biomarker measurements by utilizing the following variables in the full WHIMS dataset (N = 7479): participation in various WHI sub-studies, MCI or dementia incidence, age, region, race, ethnicity, smoking status, hormone therapy trial arm assignment, CVD, diabetes, any non-melanoma cancer, depressive symptoms, hysterectomy, history of female hormone use, BMI, and systolic blood pressure. Covariates with missing data were imputed using multivariate imputation by chained equations.^42^

#### 2.5.3 Associations of Baseline Epigenetic Age Acceleration with Longitudinal Changes in Plasma Biomarkers

To examine associations of baseline EAA with rates of change per decade in plasma biomarkers over follow-up, linear mixed effects models with random intercepts and slopes were conducted, with each plasma biomarker modeled as the dependent variable.^43,44^ An unstructured covariance matrix was specified. EAA and plasma biomarkers were similarly standardized by Z-transformation and log_2_ Z-transformation as above, respectively. Model fit comparison based on the Akaike Information Criterion indicated that linear time trend models were the most appropriate. Minimally and fully adjusted models included the same baseline covariates as described above, in addition to time from baseline to the second visit measuring plasma biomarkers. The linear mixed effects model equation was as follows:

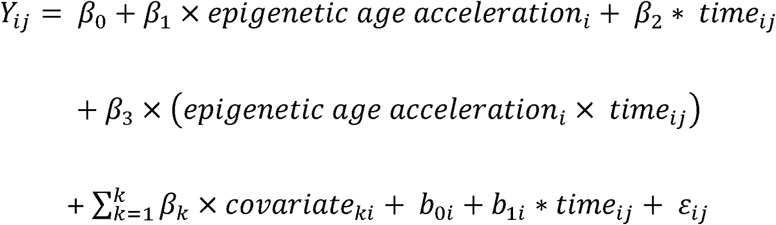

where *Y_ij_* is the outcome (standardized log_2_ plasma biomarker) for participant i at time j, *β*_0_ is the fixed intercept, *β*_1_ is the fixed effect for EAA, *β*_2_ is the fixed effect for time, *β*_3_ is the fixed effect for the interaction between EAA and time, *β*_k_ is the fixed effect of the k-th covariate, b_0i_ is the random intercept for participant i, *b_1i_* is the random slope for time for participant i, and *ε_ij_* is the residual error for participant i at time j.

#### 2.5.4 Sensitivity Analyses

Sensitivity analyses were performed to examine effect modification by race (Black and White). Participants from racial groups other than Black or White were excluded from sensitivity analyses because of insufficient sample size. We also examined potential effect modification by *APOE* ε4 carrier status. Because *APOE* ε4 carrier status was only available for White women, this sensitivity analysis was conducted among White women only. To formally test whether associations of EAA with plasma ADRD biomarkers differed by race or *APOE* ε4 carrier status, we used likelihood ratio tests to compare models with and without the corresponding interaction terms in the full analytic sample. Finally, we excluded participants with eGFR <60 mL/min/1.73 m², which is indicative of chronic kidney disease.

Analyses were conducted using R 4.4.3 (https://www.r-project.org/) in RStudio 2024.12.1 (https://cran.rstudio.com/). Because our analyses were hypothesis-driven and involved correlated plasma ADRD biomarkers, we did not apply Bonferroni correction, which would be overly conservative. We interpret findings in the context of 95% confidence intervals and present nominal p-values in the supplementary material. Thus, cautious interpretation is encouraged.

## 3 RESULTS

### 3.1 Participant Characteristics

Differences between WHIMS participants in our analytic sample relative to those not included in the sample are shown in **eTable 1**. In the analytic sample (N = 2,366), the mean (SD) baseline age was 69.8 (3.8) years; 74% were White, 17% were Black, 4.5% were Asian, 0.6% were American Indian or Alaskan Native, 0.3% were Native Hawaiian or other Pacific Islander, 2.7% were more than one race; and 6.5% were Hispanic or Latino (**Table 1**). Significant differences (p < 0.05) in covariates were observed across quartiles of DunedinPACE. Higher levels of DunedinPACE were associated with higher body mass index (BMI), a greater proportion of current or past smokers, fewer years of education, and lower physical activity. The proportion of Black and Hispanic women increased with higher levels of DunedinPACE while the proportion of White women decreased. Furthermore, higher levels of DunedinPACE were associated with higher prevalence of diabetes and hypertension and lower levels of total and high-density lipoprotein cholesterol (HDL). There were no significant differences by chronological age, cardiovascular disease (CVD), estimated glomerular filtration rate (eGFR), or apolipoprotein E epsilon 4 (*APOE* ε4) carriage.

**Table 1.**
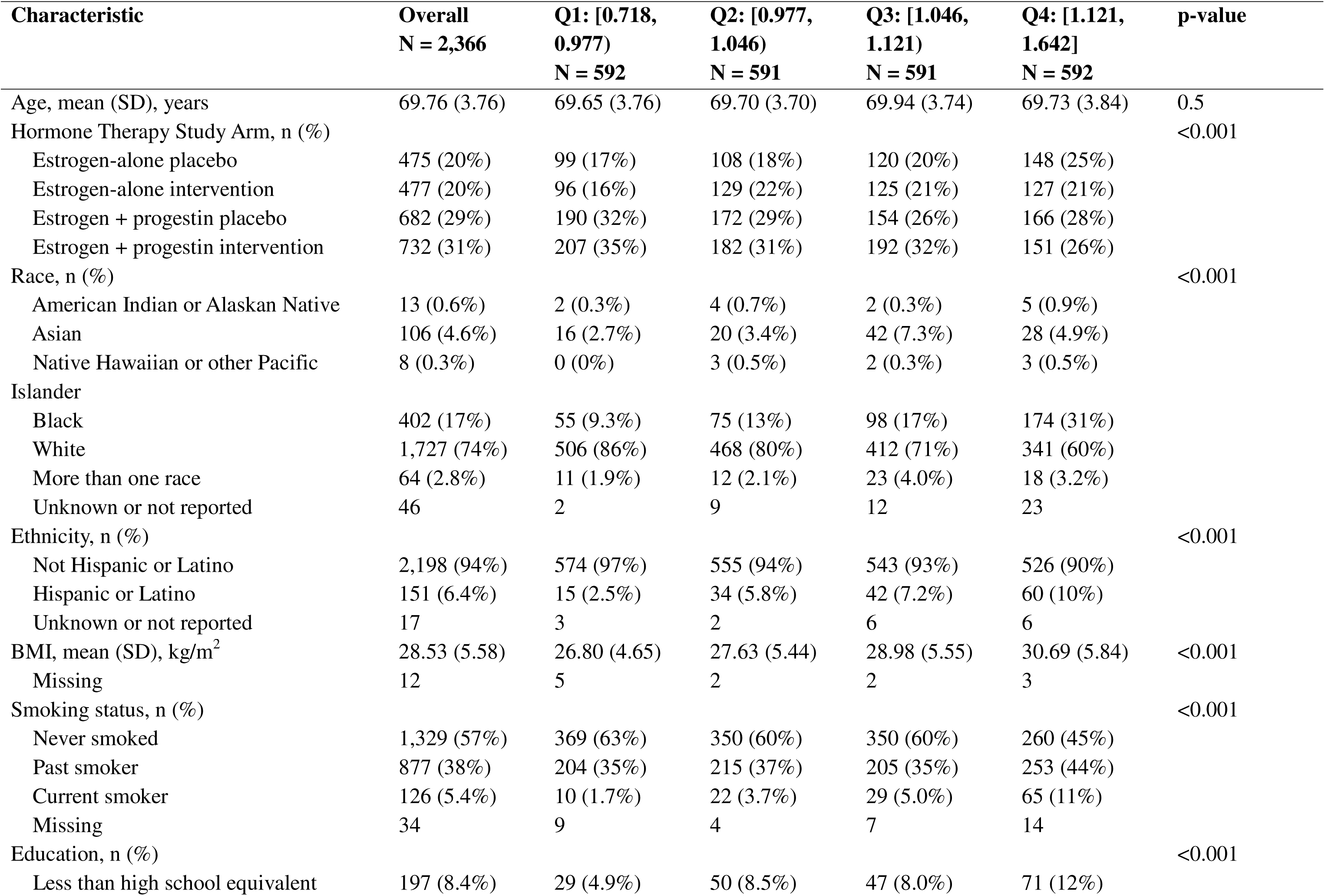
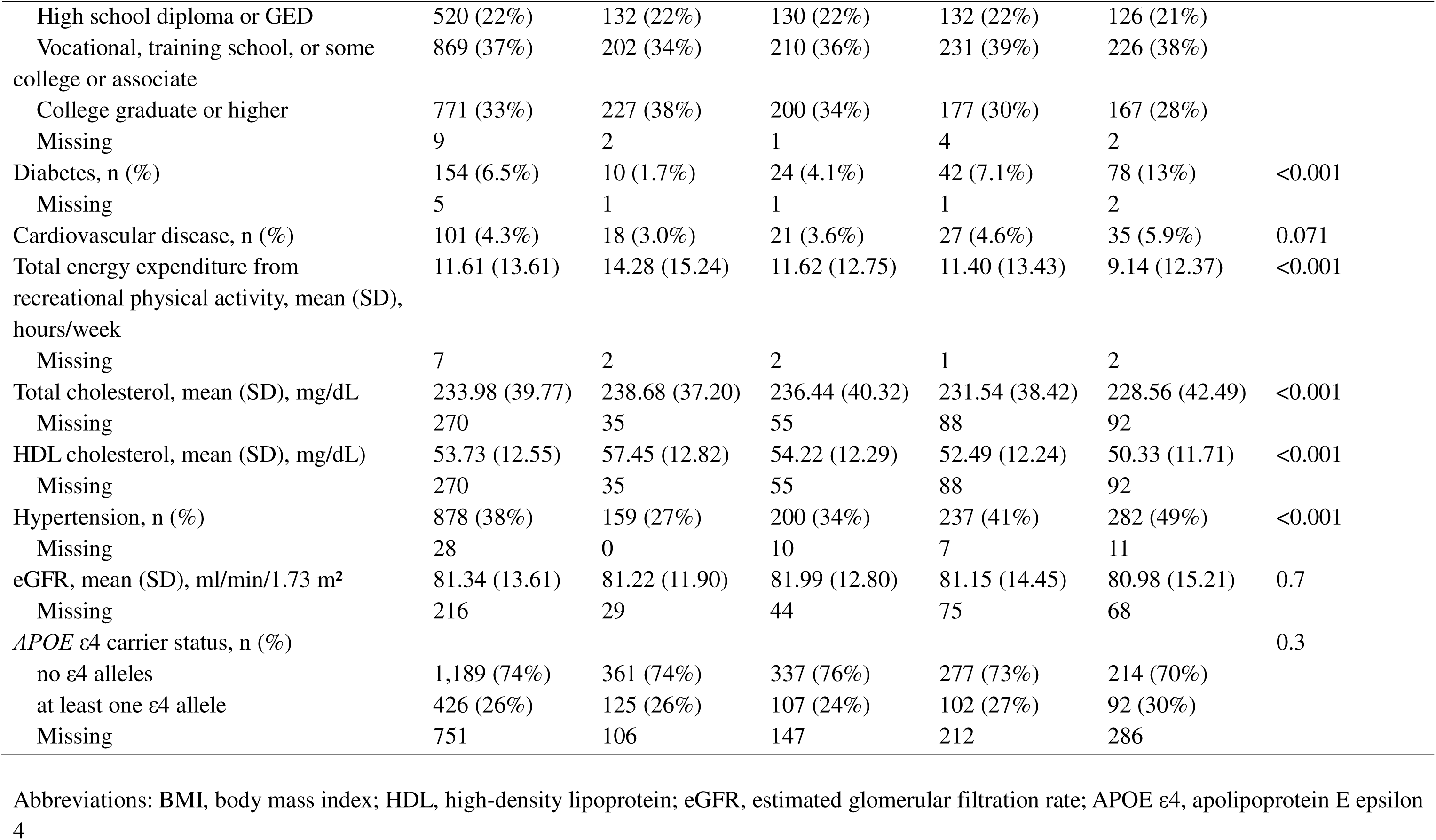
Baseline Sociodemographic, Behavior, and Health Characteristics by Quartiles of DunedinPACE.

### 3.2 Cross-Sectional Associations of Epigenetic Clocks with Plasma ADRD Biomarkers at Baseline

At baseline, most plasma biomarkers showed moderate to strong correlations (r = 0.30 to 0.69, **eFigure1**), particularly among the tau proteins (r = 0.69) and between NfL and GFAP (r = 0.51). In contrast, the Aβ42:Aβ40 ratio was moderately to weakly correlated with tau proteins (r = –0.21 to –0.34) and showed little to no correlation with NfL (r = –0.08) and GFAP (r = –0.12). Cross-sectional associations of epigenetic clocks with plasma biomarker levels at baseline are shown in **Figure 1** and **eTable 2.** In the fully-adjusted model, every one standard deviation increase in AgeAccelPheno (6.81 years) was associated with lower Aβ42:Aβ40 (β =-0.056; 95% confidence interval (CI): – 0.100 to –0.012). Associations of EAA as measured by AgeAccelHorvath, AgeAccelGrim2, and DunedinPACE with Aβ42:Aβ40 were in the same direction but failed to reach statistical significance. There were no significant associations of any epigenetic clock with p-tau181 or p-tau217. Every one standard deviation increase in AgeAccelGrim2 (4.36 years) was associated with higher NfL levels (β = 0.070, 95% CI: 0.019 to 0.120); no other clocks showed associations with NfL. There were no associations of any epigenetic clock with GFAP levels at baseline.

**Figure 1.**
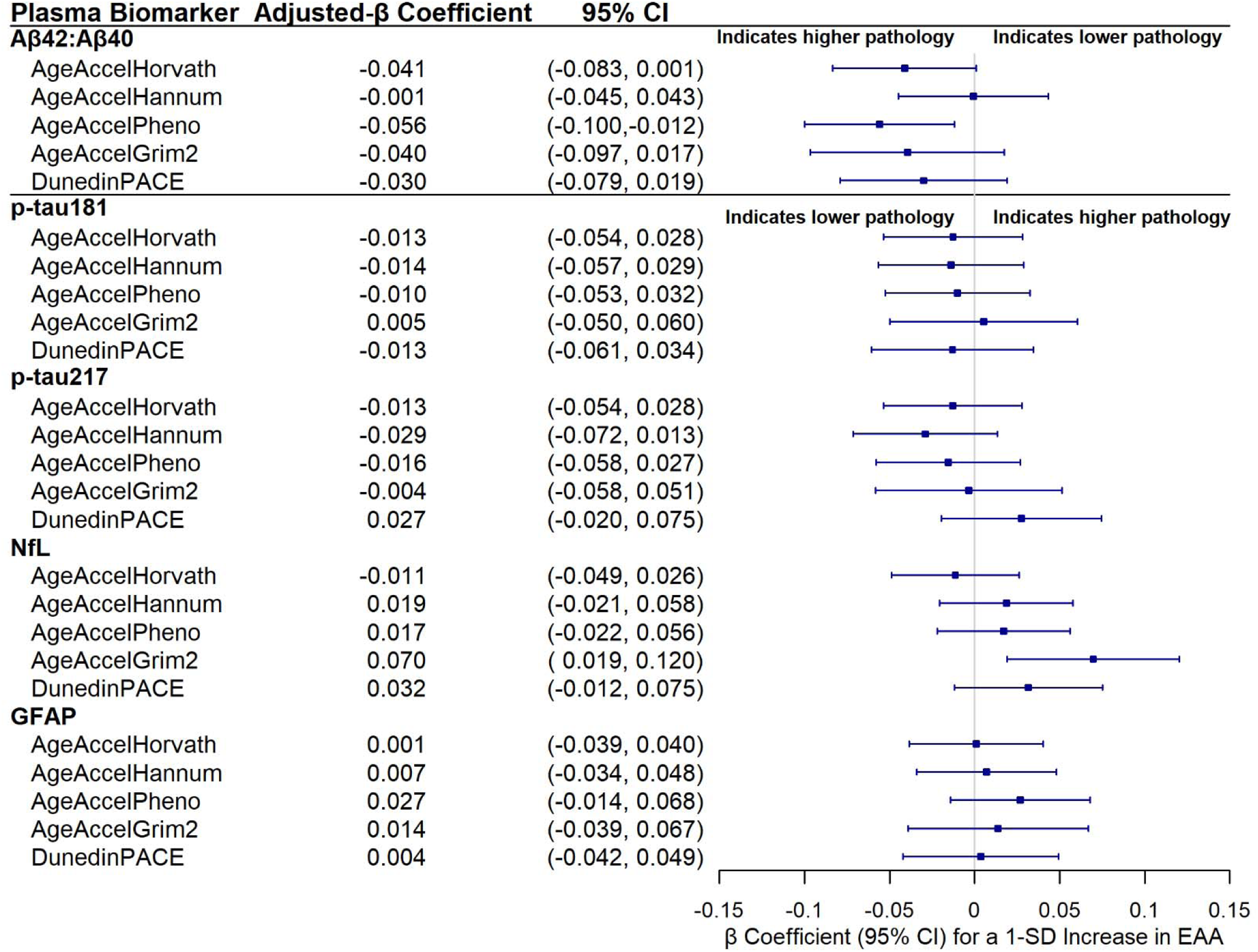
Associations of Epigenetic Clocks with Plasma Biomarkers of ADRD at Baseline. Abbreviations: EAA = Epigenetic Age Acceleration; ADRD = Alzheimer’s diseases and related dementias; CI = confidence interval; SD = standard deviation; Aβ = amyloid-β; p-tau181 = tau phosphorylated at threonine 181; p-tau217 = tau phosphorylated at threonine 217; GFAP = glial fibrillary acidic protein; NfL = neurofilament light. Associations between epigenetic age acceleration (EAA) measures and plasma biomarkers were based on linear regression models. Standardized log2-transformed biomarkers levels are shown with 95% confidence intervals per 1-SD increase in EAA. EAA include AgeAccelHorvath (SD 5.38 years), AgeAccelHannum (SD 5.01 years), AgeAccelPheno (SD 6.81 years), AgeAccelGrim2 (SD 4.36 years), and DunedinPACE (SD 0.11 years of physiologic decline). Biomarkers include Aβ42:Aβ40, GFAP, NfL, p-tau181, and p-tau217. Models were adjusted for age, hormone therapy treatment arm, education, smoking status, race, ethnicity, physical activity, body mass index, diabetes, cardiovascular disease, hypertension, total cholesterol, HDL cholesterol, estimated glomerular filtration rate, and blood cell composition.

### 3.3 Associations of Baseline Epigenetic Clocks with Longitudinal Changes in Plasma ADRD Biomarkers

Average levels of the six plasma ADRD biomarkers at baseline and an average of 15 years later are shown in **Figure 2**. The Aβ42:Aβ40 ratio did not change over time. In contrast, all other plasma biomarkers increased from baseline to the second time point 15 years later. The correlation pattern of annualized biomarker changes was similar but weaker (r = –0.03 to 0.64) compared to their correlations at baseline (e**Figure 2**).

**Figure 2.**
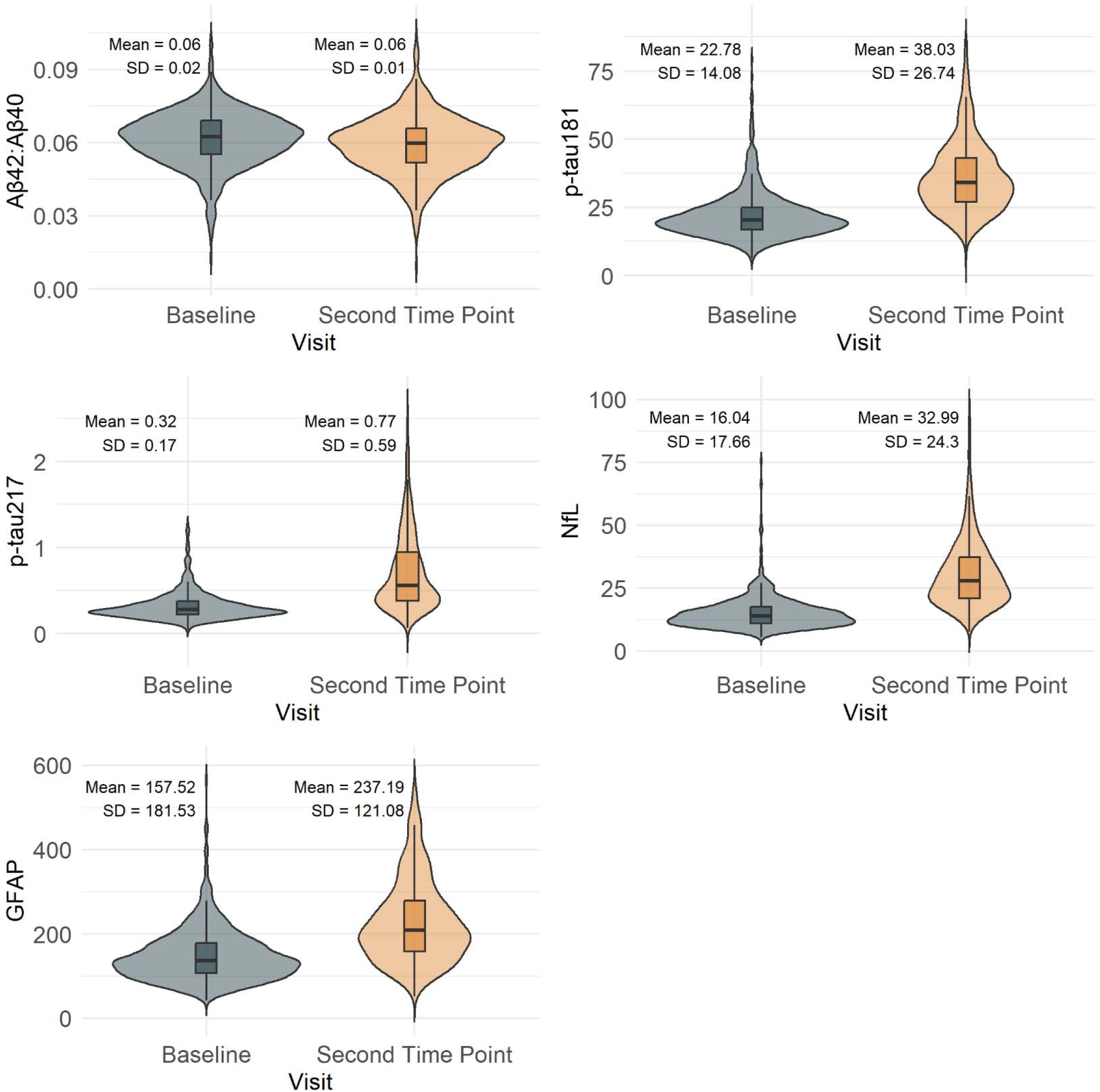
Changes in Plasma Biomarkers over an Average of 15 years. Abbreviations: SD = standard deviation; Aβ = amyloid-β; p-tau181 = tau phosphorylated at threonine 181; p-tau217 = tau phosphorylated at threonine 217; GFAP = glial fibrillary acidic protein; NfL = neurofilament light. Distribution of plasma biomarker levels at baseline and a second time point an average of 15 years later among participants with both measurements (N= 873). Biomarkers include plasma Aβ42:Aβ40, p-tau181, p-tau217, GFAP, NfL. Violin plots display the distribution density. Boxplots indicate median and interquartile ranges. Means and SDs are presented above each group. Values above the 99^th^ percentile were truncated to reduce the influence of extreme values.

Aβ42:Aβ40 changes were not correlated with the other plasma biomarkers (r = –0.03 to – 0.11), suggesting that its longitudinal trajectory was relatively independent or had a different time-course in relation to other plasma biomarkers.

Figure 3 shows associations of baseline EAA with biomarker change from fully-adjusted models. Every one standard deviation increase in DunedinPACE (indicating 0.11 years of physiological decline) was associated with a faster rate of increase in p-tau181 (β = 0.061; 95% CI = 0.022 to 0.100), p-tau217 (β = 0.056; 95% CI = 0.020 to 0.093), NfL (β = 0.062; 95% CI = 0.024 to 0.099), and GFAP (β = 0.053; 95% CI = 0.019 to 0.088) (Figure 3, Figure 4**, eTable 3**). The only other epigenetic clock that was associated with plasma biomarker change over time was AgeAccelGrim2, which was positively associated with a faster rate of increase in p-tau181 (β = 0.044; 95% CI = 0.005 to 0.083) (Figure 3).

**Figure 3.**
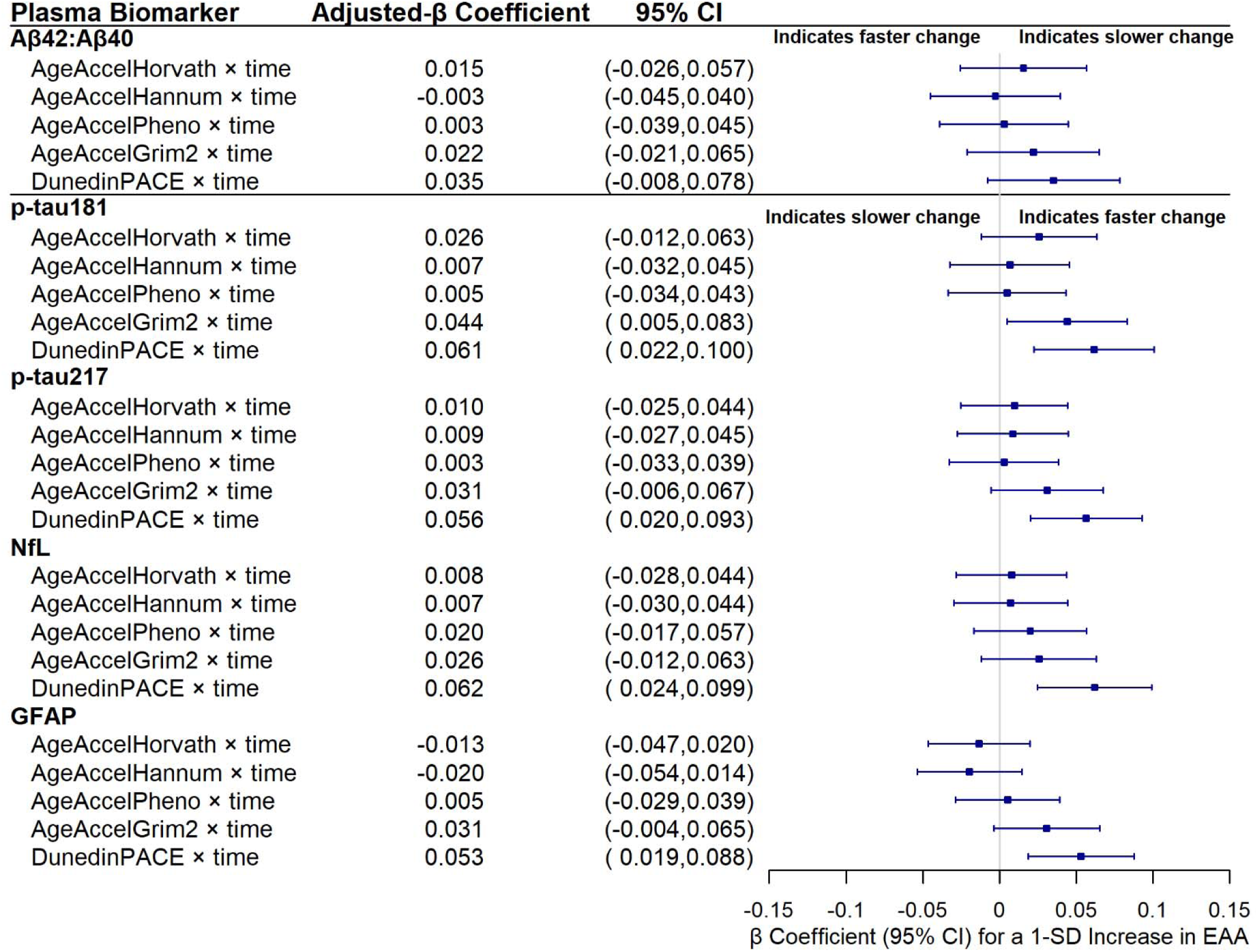
Associations of Baseline Epigenetic Clocks with Changes in Plasma Biomarkers of ADRD Over an Average of 15 Years. Abbreviations: EAA = Epigenetic Age Acceleration; ADRD = Alzheimer’s diseases and related dementias; CI = confidence intervals; SD = standard deviation; Aβ = amyloid-β; p-tau181 = tau phosphorylated at threonine 181; p-tau217 = tau phosphorylated at threonine 217; GFAP = glial fibrillary acidic protein; NfL = neurofilament light. Associations between baseline epigenetic age acceleration (EAA) measures and rates of changes per decade in plasma biomarkers based on linear mixed effects regression models. Standardized log2-transformed rate of change in plasma biomarkers are shown with 95% confidence intervals per 1-SD increase in EAA at baseline. EAA include AgeAccelHorvath (SD 5.35 years), AgeAccelHannum (SD 4.92 years), AgeAccelPheno (SD 6.73 years), AgeAccelGrim2 (SD 4.28 years), and DunedinPACE (SD 0.11 years of physiologic decline). Biomarkers include Aβ42:Aβ40, GFAP, NfL, p-tau181, and p-tau217. Models were adjusted for age, time since baseline in decades, hormone therapy treatment arm, education, smoking status, race, ethnicity, physical activity, body mass index, diabetes, cardiovascular disease, hypertension, total cholesterol, HDL cholesterol, estimated glomerular filtration rate, and blood cell composition.

**Figure 4.**
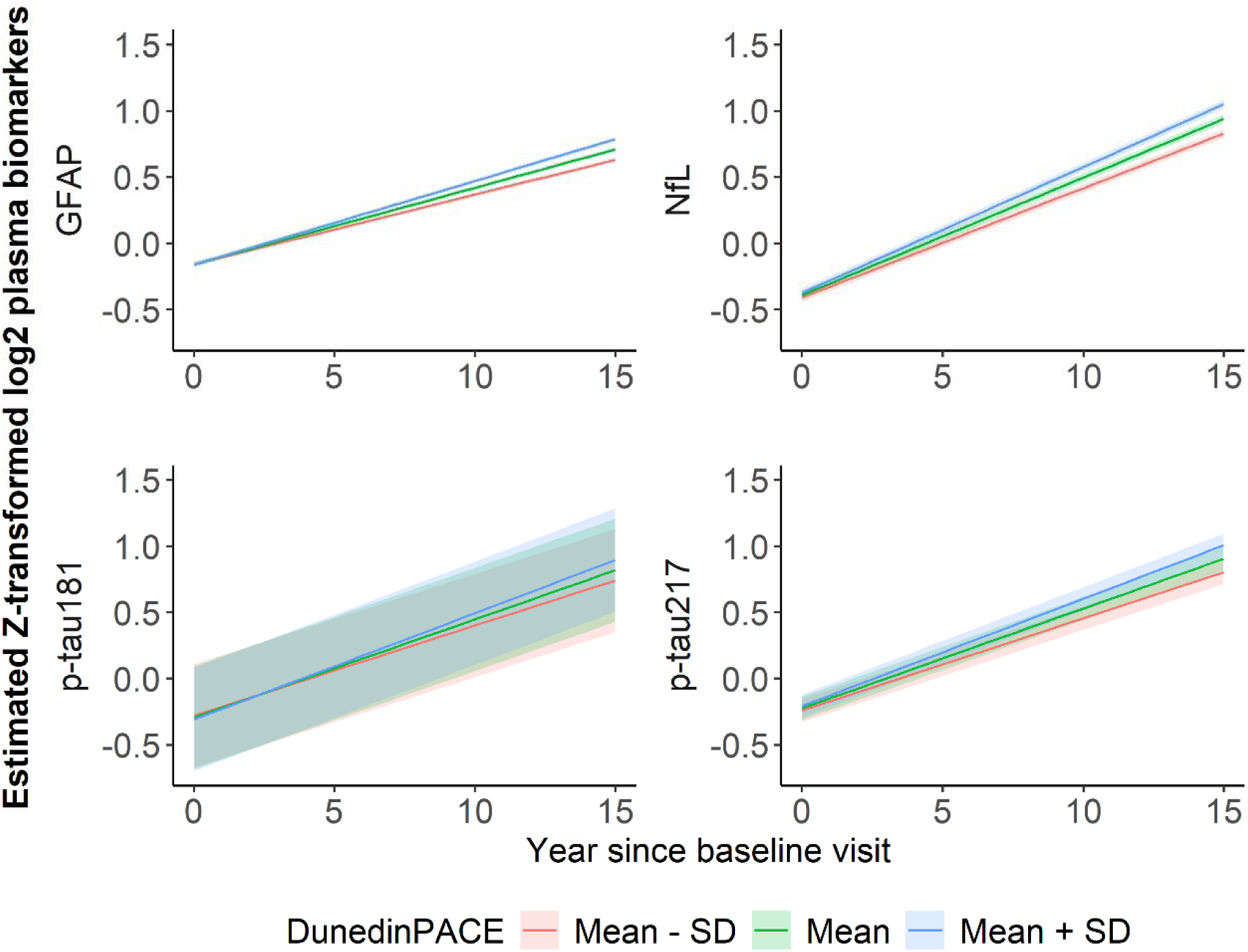
Estimated Changes in Plasma Biomarkers of ADRD over an Average of 15 Years across 3 Levels of DunedinPACE from a Fully Adjusted Linear Mixed-Effects Model. Abbreviations: ADRD = Alzheimer’s disease and related dementias; SD = standard deviation; p-tau181 = tau phosphorylated at threonine 181; p-tau217 = tau phosphorylated at threonine 217; GFAP = glial fibrillary acidic protein; NfL = neurofilament light. Linear mixed-effects models contained random intercepts and slopes adjusted for age, time since baseline, hormone therapy treatment arm, education, smoking status, race, ethnicity, physical activity, body mass index, diabetes, cardiovascular disease, hypertension, total cholesterol, HDL cholesterol, estimated glomerular filtration rate, and blood cell composition. An interaction term between DunedinPACE and time was included. Results are Z-transformed DunedinPACE set to the mean (1.05) or 1-SD below (0.94) and above the mean (1.16). Biomarkers include GFAP, NfL, p-tau181, and p-tau217.

### 3.4 Sensitivity Analyses

In the cross-sectional analysis, only two associations were modified by race: AgeAccelGrim2 with NfL (p-interaction = 0.002) and with GFAP (p-interaction = 0.034). AgeAccelGrim2 was significantly and positively associated with NfL (β = 0.183; 95% CI = 0.062 to 0.303) and GFAP (β = 0.137; 95% CI = 0.004 to 0.269) among Black women but not White women (**eTable 4**).

In the longitudinal analyses, the interaction term for race by time was significant for AgeAccelPheno and Aβ42:Aβ40 (p-interaction = 0.011), with the associations stronger but not significant among Black relative to White women (**eTable 5**). *APOE* ε4 status did not modify associations of epigenetic clocks with plasma biomarker levels at baseline or with change in plasma biomarker levels over time.

After excluding participants with eGFR < 60 mL/min/1.73 m², indicative of chronic kidney disease, 2,002 women at baseline and 832 at the second time point remained in the sensitivity analysis. At baseline, AgeAccelPheno remained associated with Aβ42:Aβ40 (p = 0.038), but AgeAccelGrim2 was no longer associated with NfL (p = 0.116; **eTable 6**). In longitudinal analyses, baseline DunedinPACE remained associated with changes in all plasma ADRD biomarkers except Aβ42:Aβ40 (**eTable 7**). Baseline AgeAccelGrim2 was no longer associated with changes in p-tau181 (p = 0.073).

## 4 DISCUSSION

Among a large, diverse cohort of women, we observed that EAA was associated with plasma biomarkers indicative of greater ADRD pathology at baseline, and with evidence of increasing plasma biomarker levels of ADRD pathology over time. Specifically, we found that EAA measured by AgeAccelPheno was associated with lower Aβ42:Aβ40 ratio, indicative of greater brain amyloid pathology, but not with other biomarkers of AD pathology at baseline (p-tau181 or p-tau217). Furthermore, EAA measured by AgeAccelGrim2 was associated with higher NfL, indicative of axonal injury, at baseline. All biomarkers except Aβ42:Aβ40 increased over the average 15-year follow-up period, indicating increasing ADRD pathology over time. In longitudinal analyses, we observed that the pace of biological aging as measured by DunedinPACE was associated with greater increases in p-tau181, p-tau217, NfL, and GFAP over time. There was limited indication of differences in associations of EAA with plasma biomarkers between White and Black women. None of the associations between EAA and biomarkers differed by *APOE* ε4 status.

Several studies have observed associations of EAA with lower cognitive performance and greater cognitive decline over time as well as increased dementia risk.^10–19,30^ Only one prior study has evaluated associations of EAA with plasma biomarkers of ADRD; however, that study was cross-sectional and did not examine p-tau217.^30^ In the Study of Latinos-Investigation of Neurocognitive Aging (SOL-INCA), GrimAge2, DunedinPACE, and principal components-based versions of several clocks (PC-Horvath, PC-Hannum, and PC-PhenoAge) were all cross-sectionally associated with plasma NfL.^30^ In contrast, we found that only AgeAccelGrim2 was associated with NfL at baseline. In agreement with our findings, PC PhenoAge was associated with lower Aβ42:Aβ40 but not p-tau181 in females.^30^ Discrepancies between WHIMS and SOL-INCA may be partly explained by differences in study populations, as SOL-INCA included only Hispanic/Latino men and women who had either MCI or were cognitively healthy, while WHIMS consisted of White, Black, and Hispanic/Latina women who were cognitively unimpaired at baseline.

In the Alzheimer’s Disease Neuroimaging Initiative, first– and second-generation clocks were not associated with cognitive functioning, but higher DunedinPACE was associated with worse scores on test of memory and executive function.^19^ On the other hand, some studies have found associations of first-generation and second-generation clocks with cognitive function.^11,14,18^ These studies, however, did not examine DunedinPACE. More recent studies found weaker or insignificant associations of DunedinPACE with cognitive decline or risk of dementia relative to first-or second-generation clocks.^12,15^ Among a subset of WHIMS women, we previously observed that higher DunedinPACE was associated with faster annual decline in cognitive function.^10^ In the larger WHIMS cohort, we found that both DunedinPACE and AgeAccelGrim2 were associated with higher risk of MCI or dementia (Nguyen et al., manuscript under review). The Framingham Heart Study reported similar findings for DunedinPACE.^13,19^

Although DunedinPACE was not associated with plasma ADRD biomarker levels at baseline, this clock was the most predictive of longitudinal change across multiple plasma biomarkers in our study. This may be partly explained by the fact that DunedinPACE was trained on longitudinal physiological change in 19 biomarkers measuring organ system integrity (e.g., cardiovascular, metabolic, renal, immune) at four time points spanning two decades, whereas other clocks were trained on chronological age (first-generation clocks) or clinical phenotypes (second-generation clocks) at a single timepoint.^8^ It is of interest that DunedinPACE was developed to predict pace of organ aging among a primarily White sample of younger adults – aged 26 to 45 years old, an age range too young to experience any substantial effect of latent neurodegenerative disorders. It is also of interest that none of the biomarkers in DunedinPACE relate specifically to brain health. Nevertheless, a measure that captures 20-year change in organ health from young adulthood to middle age significantly predicted increase in ADRD pathology over time among our diverse sample of older women, with no differences in associations between White and Black women or between those with and without the *APOE* e4 genetic risk factor. These findings support the robustness of DunedinPACE for predicting aging phenotypes and support the foundation of the geroscience hypothesis that biological mechanisms of aging may contribute to multiple co-morbidities of aging, including neurodegenerative disorders.^45^

In this study, we examined associations of EAA with changes in several plasma biomarkers associated with ADRD. These biomarkers reflect different pathologies in the brain and have different temporal dynamics in relation to dementia onset. The plasma Aβ42:Aβ40 ratio is believed to reflect soluble amyloid pathology, which develops early in the course of the disease, decades prior to dementia, and plateaus early in the preclinical period prior to the emergence of brain amyloid plaques.^46^ Furthermore, changes in plasma Aβ42:Aβ40 ratio over time are very subtle, with relative change peaking at –1% per year, and detecting such small shifts requires highly precise assays.^46,47^ In contrast, plasma ptau181 and ptau217, which are believed to reflect fibrillar amyloid pathology or amyloid plaques measured by amyloid PET, increase as AD progresses.^46,48^ In our cohort, the plasma Aβ42:Aβ40 ratio remained stable over the average 15 year follow-up, while plasma ptau181 and ptau217 increased over time, which is consistent with this dynamic. Our result showed that EAA as measured by AgeAccelPheno was associated with lower plasma Aβ42:Aβ40 ratio, but not with plasma p-tau181 or p-tau 217, at baseline.

Plasma NfL and GFAP also increased over the average 15-year follow-up in our study. Plasma NfL is a non-specific marker of axonal damage that increases with ADRD pathology.^46^ Because axonal degeneration occurs in aging and in multiple age-related pathologies, it is not surprising that NfL showed the largest increase over time in our community-based sample.^49^ Plasma GFAP, a biomarker of reactive gliosis and neuroinflammation, is also a non-specific biomarker of ADRD.^50^ Reactive gliosis is associated with amyloid plaques in the brain, and plasma GFAP is predictive of brain amyloid pathology.^51,52^ Plasma GFAP increases across the AD severity spectrum from preclinical stages to dementia.^25,29,46^ We observed moderate correlations between longitudinal plasma ptau181, ptau217, NfL and GFAP, which was expected given that they are all sensitive to increasing fibrillar amyloid pathology.^46,53,54^ DunedinPACE showed similar magnitudes of association with each of these four biomarkers in our study.

Our study’s strengths include a diverse sample with longitudinal measures of plasma ADRD biomarkers at two visits spanning an average of 15 years and examination of first-, second-, and third-generation epigenetic clocks. We had information on six plasma ADRD biomarkers, including p-tau217, which has shown better predictive performance of AD pathology and clinical phenotypes relative to other biomarkers, such as p-tau181 and p-tau231.^55,56^ The WHIMS dataset is highly comprehensive and allowed us to adjust for a robust set of potential confounders in our analysis. We also acknowledge several limitations. The sample consisted of women only, which limits the generalizability of our findings to men. Plasma ADRD biomarkers were measured at only two time points; thus, we were unable to examine biomarker trajectories over time. Because we analyzed multiple epigenetic clocks and multiple plasma biomarkers, findings should be interpreted with caution.

Accelerated pace of biological aging, as indicated by DunedinPACE, was associated with an increase in plasma ADRD biomarkers over time among older women. These findings suggest that increased pace of biological aging may be implicated in the future development of ADRD pathology. Further research is needed to determine whether DunedinPACE provides independent information in predicting ADRD beyond that provided by plasma biomarkers of ADRD and whether DunedinPACE may be useful for monitoring changes in risk of developing ADRD pathology in trials testing interventions for dementia prevention.

## Data Availability

All data produced in the present study are available upon reasonable request to the authors. Researchers can apply for access to WHIMS data through the Women's Health Initiative Publications and Presentations (P&P) Committee. Researchers must complete a paper proposal, which then must be reviewed and approved by the P&P Committee. Data access is granted once the research proposal is approved, and a data use agreement is signed. Procedures for data access are further explained here: https://www.whi.org/propose-a-paper

## ACKNOWLEDGEMENTS

We thank the following WHI investigators:

**Program Office**: (National Heart, Lung, and Blood Institute, Bethesda, Maryland) Jacques Rossouw, Jared Reis, and Candice Price

Clinical Coordinating Center: (Fred Hutchinson Cancer Center, Seattle, WA) Garnet Anderson, Ross Prentice, Andrea LaCroix, and Charles Kooperberg

**Steering Committee and Academic Centers**: (University of Alabama at Birmingham) Gretchen Wells; (Albert Einstein College of Medicine) Yasmin Mossavar-Rahmani; (University at Buffalo) Amy Millen; (University at Buffalo) Jean Wactawski-Wende; (Fred Hutchinson Cancer Center) Marian Neuhouser; (Fred Hutchinson Cancer Center) Holly Harris; (University of Massachusetts) Brian Silver; (University of North Carolina) Nora Franceschini; (Stanford Prevention Research Center) Marcia L. Stefanick; (The Ohio State University) Electra Paskett; (Wake Forest University) Mara Vitolins

## CONSENT STATEMENT

This study was performed in accordance with the ethical standards as laid down in the 1964 Declaration of Helsinki and its later amendments, and was approved by the Institutional Review Board at University of California San Diego. All participants provided written informed consent.

