## Supplementary Materials for "Epigenetic clocks and longitudinal plasma biomarkers of Alzheimer’s disease"

**Table of Contents**

**eTable1.** Baseline Sociodemographic, Behavioral, and Health Characteristics by Analytic Sample Inclusion Status

**eTable 2.** Associations of Epigenetic Clocks with Plasma ADRD Biomarkers at Baseline

**eTable 3.** Longitudinal Associations of Epigenetic Clocks with Changes in Plasma ADRD Biomarkers over 15 Years

**eTable 4.** Association of AgeAccelGrim2 with NfL and GFAP at Baseline, Stratified by Race.

**eTable 5.** Association of Baseline AgeAccelPheno with Change in Aβ42:Aβ40, Stratified by Race

**eTable 6.** Associations of Epigenetic Clocks with Plasma ADRD Biomarkers at Baseline among participants with eGFR < 60

**eTable 7.** Longitudinal Associations of Epigenetic Clocks with Changes in Plasma ADRD Biomarkers over 15 Years among participants with eGFR < 60

**eFigure 1.** Spearman Correlation Matrix of Plasma ADRD Biomarkers at Baseline

**eFigure 2.** Spearman Correlation Matrix of Annualized Changes in Plasma ADRD Biomarkers

**eTable1.** Baseline Sociodemographic, Behavioral, and Health Characteristics by Analytic Sample Inclusion Status

| Characteristic | Analytic Sample (N = 2,366) | Not in Analytic Sample (N = 5,133) | Full WHIMS (N = 7,479) | p-value |
|---|---|---|---|---|
| **Age, Mean (SD)** | 69.76 (3.76) | 70.29 (3.87) | 70.12 (3.84) | <0.001 |
| **Hormone Therapy Study Arm, n (%)** | | | | 0.088 |
| Estrogen-alone placebo | 475 (20%) | 1,009 (20%) | 1,484 (20%) | |
| Estrogen-alone intervention | 477 (20%) | 992 (19%) | 1,469 (20%) | |
| Estrogen plus progestin placebo | 682 (29%) | 1,620 (32%) | 2,302 (31%) | |
| Estrogen plus progestin intervention | 732 (31%) | 1,492 (29%) | 2,224 (30%) | |
| **Race, n (%)** | | | | <0.001 |
| American Indian or Alaskan Native | 13 (0.6%) | 4 (<0.1%) | 17 (0.2%) | |
| Asian | 106 (4.6%) | 21 (0.4%) | 127 (1.7%) | |
| Native Hawaiian or other Pacific Islander | 8 (0.3%) | 0 (0%) | 8 (0.1%) | |
| Black | 402 (17%) | 121 (2.4%) | 523 (7.1%) | |
| White | 1,727 (74%) | 4,922 (97%) | 6,649 (90%) | |
| More than one race | 64 (2.8%) | 16 (0.3%) | 80 (1.1%) | |
| Unknown or not reported | 46 | 29 | 75 | |
| **Ethnicity, n (%)** | | | | <0.001 |
| Not Hispanic or Latino | 2,198 (94%) | 5,035 (99%) | 7,233 (97%) | |
| Hispanic or Latino | 151 (6.4%) | 63 (1.2%) | 214 (2.9%) | |
| Unknown or not reported | 17 | 15 | 32 | |
| **BMI, Mean (SD)** | 28.53 (5.58) | 28.52 (5.75) | 28.52 (5.70) | 0.6 |
| Missing | 12 | 31 | 43 | |
| **Smoking Status, n (%)** | | | | <0.001 |
| Never smoked | 1,329 (57%) | 2,580 (51%) | 3,909 (53%) | |
| Past smoker | 877 (38%) | 2,053 (41%) | 2,930 (40%) | |
| Current smoker | 126 (5.4%) | 403 (8.0%) | 529 (7.2%) | |
| Missing | 34 | 77 | 111 | |
| **Education, n (%)** | | | | <0.001 |
| Less than high school equivalent | 197 (8.4%) | 379 (7.4%) | 576 (7.7%) | |
| High school diploma or GED | 520 (22%) | 1,127 (22%) | 1,647 (22%) | |
| Vocational, training school, or some college or associate | 869 (37%) | 2,133 (42%) | 3,002 (40%) | |
| College graduate or higher | 771 (33%) | 1,461 (29%) | 2,232 (30%) | |
| Missing | 9 | 13 | 22 | |
| **Diabetes, n (%)** | 154 (6.5%) | 334 (6.5%) | 488 (6.5%) | >0.9 |
| Missing | 5 | 9 | 14 | |
| **Cardiovascular disease, n (%)** | 101 (4.3%) | 265 (5.2%) | 366 (4.9%) | 0.088 |
| **Physical activity (hours/week), Mean (SD)** | 11.61 (13.61) | 11.09 (13.13) | 11.25 (13.29) | 0.074 |
| Missing | 7 | 10 | 17 | |
| **Total cholesterol (mg/dL), Mean (SD)** | 233.98 (39.77) | 234.57 (40.15) | 234.38 (40.03) | 0.7 |
| Missing | 270 | 715 | 985 | |
| **HDL cholesterol (mg/dL), Mean (SD)** | 53.73 (12.55) | 53.23 (12.55) | 53.40 (12.55) | 0.1 |
| Missing | 270 | 715 | 985 | |

| Characteristic | Analytic Sample (N = 2,366) | Not in Analytic Sample (N = 5,133) | Full WHIMS (N = 7,479) | p-value |
|---|---|---|---|---|
| **Hypertension, n (%)** | 878 (38%) | 2,038 (40%) | 2,916 (39%) | 0.024 |
| Missing | 28 | 58 | 86 | |
| **eGFR (ml/min/1.73 m²), Mean (SD)** | 81.34 (13.61) | 79.87 (13.35) | 80.35 (13.45) | <0.001 |
| Missing | 216 | 645 | 861 | |
| **APOE ε4 carrier status, n (%)** | | | | 0.2 |
| No ε4 alleles | 1,189 (74%) | 3,194 (75%) | 4,383 (75%) | |
| At least one ε4 allele | 426 (26%) | 1,054 (25%) | 1,480 (25%) | |
| Missing | 751 | 865 | 1,616 | |

Abbreviations: WHIMS = Women's Health Initiative Memory Study; SD = standard deviation; HDL, high-density lipoprotein; eGFR = estimated glomerular filtration rate; APOE ε4 = apolipoprotein E epsilon 4.

**eTable 2.** Associations of Epigenetic Clocks with Plasma ADRD Biomarkers at Baseline

| EAA Measure | Mean (SD) | Model | Aβ42:Aβ40 | | p-tau181 | | p-tau217 | | NfL | | GFAP | |
|---|---|---|---|---|---|---|---|---|---|---|---|---|
| | | | Adjusted-β (95% CI) | p-value | Adjusted-β (95% CI) | p-value | Adjusted-β (95% CI) | p-value | Adjusted-β (95% CI) | p-value | Adjusted-β (95% CI) | p-value |
| **AgeAccelHorvath** | -1.44 (5.38) | Model 1 | -0.035 (-0.076, 0.006) | 0.09 | -0.014 (-0.054, 0.026) | 0.486 | -0.017 (-0.057, 0.022) | 0.387 | -0.008 (-0.046, 0.030) | 0.691 | -0.009 (-0.047, 0.030) | 0.653 |
| | | Model 2 | -0.041 (-0.083, 0.001) | 0.056 | -0.013 (-0.054, 0.028) | 0.54 | -0.013 (-0.054, 0.028) | 0.53 | -0.011 (-0.049, 0.026) | 0.551 | 0.001 (-0.039, 0.040) | 0.969 |
| **AgeAccelHannum** | -1.33 (5.01) | Model 1 | 0.007 (-0.034, 0.048) | 0.729 | -0.010 (-0.050, 0.030) | 0.618 | -0.021 (-0.060, 0.019) | 0.307 | 0.036 (-0.003, 0.074) | 0.069 | 0.005 (-0.033, 0.044) | 0.791 |
| | | Model 2 | -0.001 (-0.045, 0.043) | 0.971 | -0.014 (-0.057, 0.029) | 0.52 | -0.029 (-0.072, 0.013) | 0.179 | 0.019 (-0.021, 0.058) | 0.35 | 0.007 (-0.034, 0.048) | 0.745 |
| **AgeAccelPheno** | -1.65 (6.81) | Model 1 | -0.050 (-0.091, -0.009) | **0.016** | 0.005 (-0.035, 0.045) | 0.794 | -0.002 (-0.042, 0.037) | 0.911 | 0.036 (-0.002, 0.074) | 0.064 | 0.011 (-0.027, 0.050) | 0.566 |
| | | Model 2 | -0.056 (-0.100, -0.012) | **0.013** | -0.010 (-0.053, 0.032) | 0.638 | -0.016 (-0.058, 0.027) | 0.473 | 0.017 (-0.022, 0.056) | 0.395 | 0.027 (-0.014, 0.068) | 0.2 |
| **AgeAccelGrim2** | -1.01 (4.36) | Model 1 | -0.033 (-0.081, 0.014) | 0.166 | 0.026 (-0.020, 0.072) | 0.272 | 0.018 (-0.028, 0.064) | 0.446 | 0.081 (0.037, 0.125) | **< 0.001** | -0.021 (-0.065, 0.024) | 0.36 |
| | | Model 2 | -0.040 (-0.097, 0.017) | 0.173 | 0.005 (-0.050, 0.060) | 0.849 | -0.004 (-0.058, 0.051) | 0.898 | 0.070 (0.019, 0.120) | **0.007** | 0.014 (-0.039, 0.067) | 0.611 |
| **DunedinPACE** | 1.05 (0.11) | Model 1 | -0.041 (-0.084, 0.003) | 0.067 | 0.006 (-0.036, 0.049) | 0.778 | 0.025 (-0.017, 0.067) | 0.243 | 0.022 (-0.018, 0.063) | 0.278 | -0.045 (-0.086, -0.004) | **0.03** |
| | | Model 2 | -0.030 (-0.079, 0.019) | 0.229 | -0.013 (-0.061, 0.034) | 0.588 | 0.027 (-0.020, 0.075) | 0.254 | 0.032 (-0.012, 0.075) | 0.155 | 0.004 (-0.042, 0.049) | 0.879 |

Abbreviations: ADRD = Alzheimer's disease and related dementias; EAA = Epigenetic Age Acceleration; SD = standard deviation; CI = Confidence Interval; Aβ = amyloid-β; p-tau181 = tau phosphorylated at threonine 181; p-tau217 = tau phosphorylated at threonine 217; GFAP = glial fibrillary acidic protein; NfL = neurofilament light.

Bold indicates a significant 95% confidence interval ($p < 0.05$). Model 1 adjusted for chronological age, hormone therapy treatment arm, education, smoking, race, and ethnicity. Model 2 additionally adjusted for physical activity, body mass index, diabetes, cardiovascular disease, hypertension, total cholesterol, HDL cholesterol, estimated glomerular filtration rate, and blood cell composition.

Sample Size: 2304; participants with missing race or ethnicity were excluded.

**eTable 3.** Longitudinal Associations of Epigenetic Clocks with Changes in Plasma ADRD Biomarkers over 15 Years

| EAA × Time Interaction | EAA Mean (SD) | Model | Aβ42:Aβ40 | | p-tau181 | | p-tau217 | | NfL | | GFAP | |
|---|---|---|---|---|---|---|---|---|---|---|---|---|
| | | | Adjusted-β (95% CI) | p-value | Adjusted-β (95% CI) | p-value | Adjusted-β (95% CI) | p-value | Adjusted-β (95% CI) | p-value | Adjusted-β (95% CI) | p-value |
| **AgeAccelHorvath × time** | -1.46 (5.35) | Model 1 | 0.015 (-0.026, 0.056) | 0.467 | 0.026 (-0.012, 0.064) | 0.18 | 0.010 (-0.025, 0.044) | 0.588 | 0.006 (-0.030, 0.042) | 0.734 | -0.014 (-0.048, 0.019) | 0.393 |
| | | Model 2 | 0.015 (-0.026, 0.057) | 0.462 | 0.026 (-0.012, 0.063) | 0.182 | 0.010 (-0.025, 0.044) | 0.592 | 0.008 (-0.028, 0.044) | 0.674 | -0.013 (-0.047, 0.020) | 0.427 |
| **AgeAccelHannum × time** | -1.39 (4.92) | Model 1 | -0.003 (-0.045, 0.039) | 0.892 | 0.006 (-0.033, 0.045) | 0.76 | 0.009 (-0.027, 0.045) | 0.628 | 0.005 (-0.032, 0.042) | 0.796 | -0.020 (-0.054, 0.014) | 0.25 |
| | | Model 2 | -0.003 (-0.045, 0.040) | 0.898 | 0.007 (-0.032, 0.045) | 0.741 | 0.009 (-0.027, 0.045) | 0.638 | 0.007 (-0.030, 0.044) | 0.705 | -0.020 (-0.054, 0.014) | 0.261 |
| **AgeAccelPheno × time** | -1.83 (6.73) | Model 1 | 0.003 (-0.039, 0.044) | 0.907 | 0.003 (-0.035, 0.042) | 0.866 | 0.002 (-0.033, 0.038) | 0.896 | 0.015 (-0.022, 0.051) | 0.429 | 0.003 (-0.030, 0.037) | 0.846 |
| | | Model 2 | 0.003 (-0.039, 0.045) | 0.893 | 0.005 (-0.034, 0.043) | 0.808 | 0.003 (-0.033, 0.039) | 0.876 | 0.020 (-0.017, 0.057) | 0.289 | 0.005 (-0.029, 0.039) | 0.768 |
| **AgeAccelGrim2 × time** | -1.20 (4.28) | Model 1 | 0.023 (-0.020, 0.065) | 0.303 | 0.039 (0.000, 0.078) | 0.051 | 0.030 (-0.007, 0.066) | 0.113 | 0.016 (-0.021, 0.054) | 0.393 | 0.028 (-0.007, 0.062) | 0.115 |
| | | Model 2 | 0.022 (-0.021, 0.065) | 0.32 | 0.044 (0.005, 0.083) | **0.029** | 0.031 (-0.006, 0.067) | 0.099 | 0.026 (-0.012, 0.063) | 0.181 | 0.031 (-0.004, 0.065) | 0.083 |
| **DunedinPACE × time** | 1.05 (0.11) | Model 1 | 0.035 (-0.008, 0.078) | 0.114 | 0.055 (0.016, 0.094) | **0.006** | 0.055 (0.019, 0.091) | **0.003** | 0.052 (0.015, 0.090) | **0.006** | 0.051 (0.016, 0.085) | **0.004** |
| | | Model 2 | 0.035 (-0.008, 0.078) | 0.111 | 0.061 (0.022, 0.100) | **0.002** | 0.056 (0.020, 0.093) | **0.003** | 0.062 (0.024, 0.099) | **0.001** | 0.053 (0.019, 0.088) | **0.003** |

Abbreviations: ADRD = Alzheimer's disease and related dementias; EAA = Epigenetic Age Acceleration; SD = standard deviation; CI = Confidence Interval; Aβ = amyloid beta; p-tau181 = tau phosphorylated at threonine 181; p-tau217 = tau phosphorylated at threonine 217; GFAP = glial fibrillary acidic protein; NfL = neurofilament light.

Bold indicates significant 95% confidence interval ($p < 0.05$). Model 1 adjusted for chronological age, time since baseline, hormone therapy treatment arm, education, smoking, race, ethnicity. Model 2 additional adjusted for physical activity, body mass index, diabetes, cardiovascular disease, hypertension, total cholesterol, HDL cholesterol, estimated glomerular filtration rate, blood cell composition.

Sample Size at second time point: 873

**eTable 4.** Association of AgeAccelGrim2 with NfL and GFAP at Baseline, Stratified by Race.

| EAA Measure | Biomarker | Model | Adjusted-β (95% CI) | | Interaction p-value |
|---|---|---|---|---|---|
| | | | **White Women** | **Black Women** | |
| AgeAccelGrim2 | NfL | Model 1 | 0.032 (–0.017, 0.081) | 0.265 (0.159, 0.370) | <0.001 |
| | | Model 2 | 0.046 (–0.011, 0.103) | **0.183 (0.062, 0.303)** | **0.002** |
| | GFAP | Model 1 | –0.048 (–0.097, 0.001) | 0.096 (–0.013, 0.205) | **0.014** |
| | | Model 2 | –0.012 (–0.072, 0.047) | **0.137 (0.004, 0.269)** | **0.034** |

Abbreviations: EAA = Epigenetic Age Acceleration; CI = Confidence Interval; GFAP = glial fibrillary acidic protein; NfL = neurofilament light.

Model 1 adjusted for chronological age, hormone therapy treatment arm, education, smoking.
Model 2 additional adjusted for physical activity, body mass index, diabetes, cardiovascular disease, Hypertension, total cholesterol, HDL cholesterol, estimated glomerular filtration rate, blood cell composition.

Interaction p-value: From likelihood ratio tests comparing nested models with and without the EAA × race interaction term.

**eTable 5.** Association of Baseline AgeAccelPheno with Change in Aβ42:Aβ40, Stratified by Race

| EAA × Time Interaction | Biomarker | Model | Adjusted-β (95% CI) | | Interaction p-value |
|---|---|---|---|---|---|
| | | | White Women | Black Women | |
| **AgeAccelPheno × time** | Aβ42:Aβ40 | Model 1 | 0.006 (–0.039, 0.050) | 0.026 (–0.138, 0.190) | **0.013** |
| | | Model 2 | 0.005 (–0.039, 0.050) | 0.049 (–0.122, 0.219) | **0.011** |

Abbreviations: EAA = Epigenetic Age Acceleration; CI = Confidence Interval; Aβ = amyloid beta

Model 1 adjusted for chronological age, time since baseline, hormone therapy treatment arm, education, smoking, race, ethnicity.
Model 2 additional adjusted for physical activity, body mass index, diabetes, cardiovascular disease, Hypertension, total cholesterol, HDL cholesterol, estimated glomerular filtration rate, blood cell composition.
Interaction p-value: From a likelihood ratio test comparing nested models with and without the EAA × time × race interaction term.

**eTable 6.** Associations of Epigenetic Clocks with Plasma ADRD Biomarkers at Baseline among participants with eGFR < 60

| EAA Measure | Mean (SD) | Model | Aβ42:Aβ40 | | p-tau181 | | p-tau217 | | NfL | | GFAP | |
|---|---|---|---|---|---|---|---|---|---|---|---|---|
| | | | Adjusted-β (95% CI) | p-value | Adjusted-β (95% CI) | p-value | Adjusted-β (95% CI) | p-value | Adjusted-β (95% CI) | p-value | Adjusted-β (95% CI) | p-value |
| **AgeAccelHorvath** | -1.42 (5.38) | Model 1 | -0.028 (-0.072, 0.017) | 0.226 | -0.028 (-0.072, 0.015) | 0.201 | -0.032 (-0.075, 0.011) | 0.146 | -0.027 (-0.069, 0.014) | 0.194 | -0.013 (-0.055, 0.028) | 0.526 |
| | | Model 2 | -0.035 (-0.081, 0.011) | 0.134 | -0.022 (-0.067, 0.022) | 0.329 | -0.023 (-0.067, 0.021) | 0.305 | -0.028 (-0.069, 0.014) | 0.189 | -0.003 (-0.045, 0.040) | 0.903 |
| **AgeAccelHannum** | -1.60 (4.85) | Model 1 | 0.013 (-0.030, 0.057) | 0.547 | -0.030 (-0.073, 0.013) | 0.172 | -0.044 (-0.086, -0.001) | **0.043** | 0.014 (-0.027, 0.055) | 0.509 | -0.012 (-0.053, 0.029) | 0.574 |
| | | Model 2 | 0.004 (-0.043, 0.051) | 0.874 | -0.027 (-0.073, 0.019) | 0.249 | -0.044 (-0.089, 0.002) | 0.059 | 0.007 (-0.036, 0.049) | 0.754 | -0.005 (-0.049, 0.038) | 0.81 |
| **AgeAccelPheno** | -2.02 (6.71) | Model 1 | -0.043 (-0.087, 0.001) | 0.056 | -0.013 (-0.056, 0.031) | 0.566 | -0.018 (-0.061, 0.024) | 0.405 | 0.001 (-0.040, 0.043) | 0.946 | -0.010 (-0.052, 0.031) | 0.623 |
| | | Model 2 | -0.050 (-0.097, -0.003) | **0.038** | -0.016 (-0.062, 0.030) | 0.485 | -0.018 (-0.063, 0.028) | 0.449 | 0.001 (-0.042, 0.044) | 0.967 | 0.013 (-0.031, 0.057) | 0.56 |
| **DunedinPACE** | 1.05 (0.11) | Model 1 | -0.034 (-0.081, 0.013) | 0.159 | -0.015 (-0.060, 0.031) | 0.535 | 0.006 (-0.040, 0.051) | 0.808 | -0.018 (-0.062, 0.026) | 0.422 | -0.062 (-0.105, -0.018) | **0.006** |
| | | Model 2 | -0.021 (-0.074, 0.032) | 0.44 | -0.024 (-0.075, 0.028) | 0.367 | 0.021 (-0.031, 0.072) | 0.429 | 0.012 (-0.036, 0.060) | 0.635 | -0.004 (-0.053, 0.046) | 0.881 |
| **AgeAccelGrim2** | -1.19 (4.26) | Model 1 | -0.026 (-0.077, 0.025) | 0.323 | 0.006 (-0.044, 0.056) | 0.811 | -0.010 (-0.059, 0.039) | 0.697 | 0.032 (-0.016, 0.079) | 0.194 | -0.049 (-0.097, -0.001) | **0.044** |
| | | Model 2 | -0.031 (-0.093, 0.031) | 0.328 | 0.002 (-0.058, 0.063) | 0.937 | -0.016 (-0.076, 0.044) | 0.598 | 0.045 (-0.011, 0.100) | 0.116 | -0.006 (-0.063, 0.052) | 0.847 |

Abbreviations: ADRD = Alzheimer's disease and related dementias; SD = standard deviation; eGFR = estimated glomerular filtration rate; EAA = Epigenetic Age Acceleration; SD = standard deviation; CI = Confidence Interval; Aβ = amyloid beta; p-tau181 = tau phosphorylated at threonine 181; p-tau217 = tau phosphorylated at threonine 217; GFAP = glial fibrillary acidic protein; NfL = neurofilament light.

Bold indicates a significant 95% confidence interval ($p < 0.05$).
Model 1 adjusted for chronological age, hormone therapy treatment arm, education, smoking, race, and ethnicity.
Model 2 additionally adjusted for physical activity, body mass index, diabetes, cardiovascular disease, hypertension, total cholesterol, HDL cholesterol, estimated glomerular filtration rate, and blood cell composition.
Sample Size: 2,002; participants with missing race, ethnicity, or eGFR <60 were excluded

**eTable 7.** Longitudinal Associations of Epigenetic Clocks with Changes in Plasma ADRD Biomarkers over 15 Years among Participants with eGFR < 60

| **EAA Measure × Time** | **Mean (SD)** | **Model** | **Aβ42:Aβ40** | | **p-tau181** | | **p-tau217** | | **NfL** | | **GFAP** | |
|---|---|---|---|---|---|---|---|---|---|---|---|---|
| | | | Adjusted-β (95% CI) | p-value | Adjusted-β (95% CI) | p-value | Adjusted-β (95% CI) | p-value | Adjusted-β (95% CI) | p-value | Adjusted-β (95% CI) | p-value |
| AgeAccelHorvath × time | -1.45 (5.35) | Model 1 | 0.014 (-0.028, 0.057) | 0.508 | 0.028 (-0.010, 0.067) | 0.153 | 0.009 (-0.027, 0.044) | 0.639 | 0.006 (-0.031, 0.043) | 0.745 | -0.014 (-0.048, 0.019) | 0.402 |
| | | Model 2 | 0.015 (-0.028, 0.057) | 0.505 | 0.028 (-0.011, 0.066) | 0.16 | 0.008 (-0.027, 0.044) | 0.643 | 0.007 (-0.030, 0.044) | 0.714 | -0.013 (-0.047, 0.021) | 0.444 |
| AgeAccelHannum × time | -1.60 (4.80) | Model 1 | -0.004 (-0.047, 0.039) | 0.862 | 0.009 (-0.030, 0.048) | 0.638 | 0.010 (-0.026, 0.046) | 0.576 | 0.005 (-0.032, 0.042) | 0.794 | -0.017 (-0.051, 0.017) | 0.334 |
| | | Model 2 | -0.004 (-0.047, 0.039) | 0.852 | 0.010 (-0.029, 0.049) | 0.631 | 0.010 (-0.026, 0.046) | 0.58 | 0.007 (-0.030, 0.044) | 0.721 | -0.017 (-0.051, 0.017) | 0.341 |
| AgeAccelPheno × time | -2.13 (6.65) | Model 1 | 0.008 (-0.035, 0.051) | 0.719 | 0.004 (-0.035, 0.043) | 0.856 | -0.004 (-0.040, 0.032) | 0.83 | 0.019 (-0.018, 0.056) | 0.309 | 0.003 (-0.031, 0.037) | 0.873 |
| | | Model 2 | 0.008 (-0.036, 0.051) | 0.732 | 0.004 (-0.035, 0.043) | 0.828 | -0.004 (-0.040, 0.032) | 0.839 | 0.023 (-0.014, 0.060) | 0.232 | 0.004 (-0.031, 0.038) | 0.83 |
| AgeAccelGrim2 × time | -1.38 (4.19) | Model 1 | 0.021 (-0.023, 0.066) | 0.341 | 0.033 (-0.006, 0.073) | 0.102 | 0.021 (-0.016, 0.058) | 0.263 | 0.028 (-0.010, 0.066) | 0.152 | 0.027 (-0.008, 0.062) | 0.136 |
| | | Model 2 | 0.022 (-0.022, 0.066) | 0.332 | 0.037 (-0.003, 0.076) | 0.073 | 0.022 (-0.015, 0.059) | 0.252 | 0.035 (-0.004, 0.073) | 0.077 | 0.028 (-0.007, 0.063) | 0.116 |
| DunedinPACE × time | 1.04 (0.11) | Model 1 | 0.033 (-0.012, 0.077) | 0.15 | 0.050 (0.010, 0.089) | **0.015** | 0.048 (0.011, 0.085) | **0.011** | 0.064 (0.027, 0.102) | **0.001** | 0.048 (0.013, 0.083) | **0.007** |
| | | Model 2 | 0.033 (-0.011, 0.078) | 0.142 | 0.054 (0.015, 0.094) | **0.008** | 0.049 (0.012, 0.086) | **0.011** | 0.071 (0.033, 0.109) | **< 0.001** | 0.049 (0.014, 0.084) | **0.007** |

Abbreviations: ADRD = Alzheimer's disease and related dementias; eGFR = estimated glomerular filtration rate; EAA = Epigenetic Age Acceleration; SD = standard deviation; CI = Confidence Interval; Aβ = amyloid beta; p-tau181 = tau phosphorylated at threonine 181; p-tau217 = tau phosphorylated at threonine 217; GFAP = glial fibrillary acidic protein; NfL = neurofilament light.

Bold indicates a significant 95% confidence interval (p < 0.05).
Model 1 adjusted for chronological age, time since baseline, hormone therapy treatment arm, education, smoking, race, ethnicity.
Model 2 additional adjusted for physical activity, body mass index, diabetes, cardiovascular disease, hypertension, total cholesterol, HDL cholesterol, estimated glomerular filtration rate, blood cell composition.
Sample Size at second time point: 832.

**eFigure 1.** Spearman Correlation Matrix of Plasma ADRD Biomarkers at Baseline

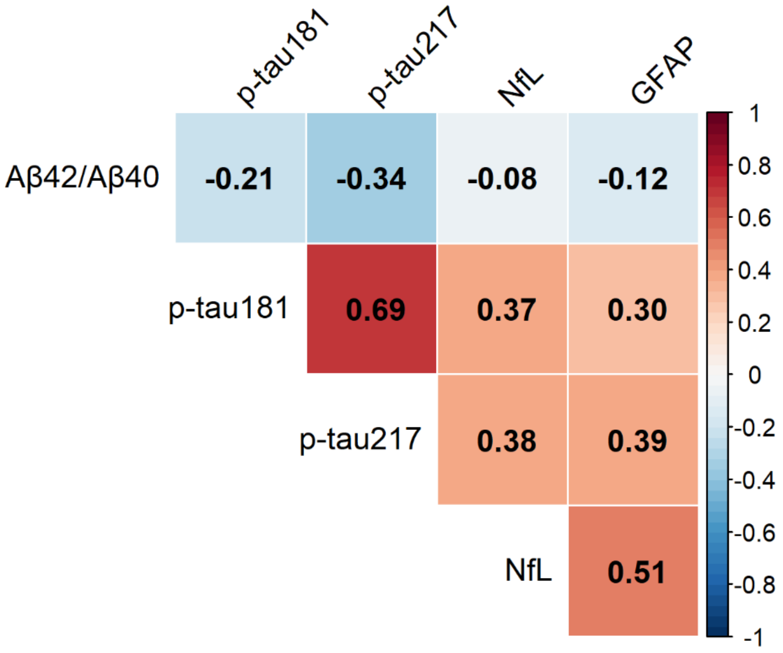


Abbreviations: ADRD = Alzheimer's disease and related dementias; Aβ = amyloid beta; p-tau181 = tau phosphorylated at threonine 181; p-tau217 = tau phosphorylated at threonine 217; GFAP = glial fibrillary acidic protein; NfL = neurofilament light.

**eFigure 2.** Spearman Correlation Matrix of Annualized Changes in Plasma ADRD Biomarkers

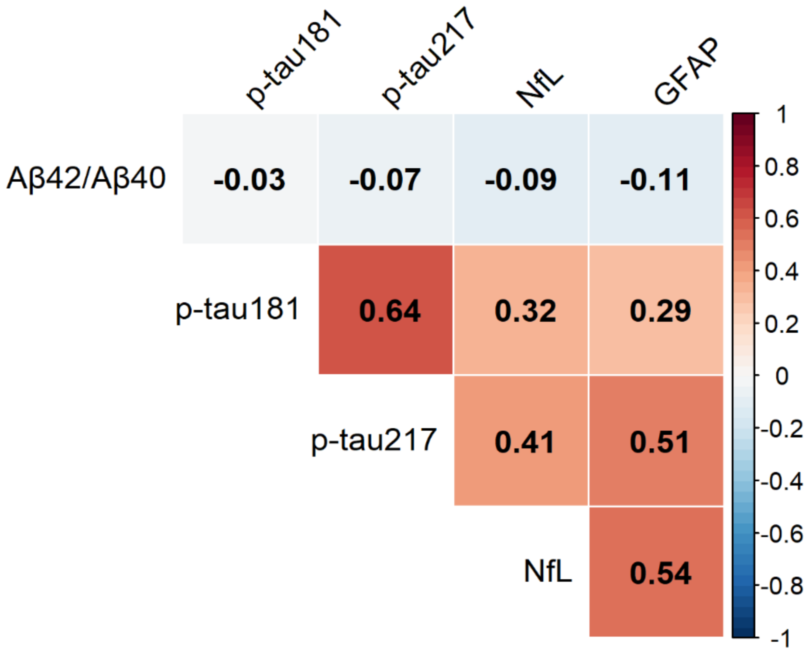


Abbreviations: ADRD = Alzheimer's disease and related dementias; Aβ = amyloid beta; p-tau181 = tau phosphorylated at threonine 181; p-tau217 = tau phosphorylated at threonine 217; GFAP = glial fibrillary acidic protein; NfL = neurofilament light.
