## Supplementary Methods for "Epigenetic clocks and longitudinal plasma biomarkers of Alzheimer’s disease"

**Methylation Quality Control**

Samples were assessed for quality using 17 control metrics from Illumina. Samples were removed if they did not meet Illumina's recommended thresholds for each control metric (N=14 removed). Sex was determined by clustering samples on the average intensity values of CpG sites on the X and Y chromosomes.[1] Samples that fell outside of the female cluster or had a mismatch between reported and detected sex were excluded (N=8 removed). Samples with mean bisulfite intensity values < 4,000 were excluded (N=1 removed). Detection p-values were calculated using out-of-band (OOB) probes.[2] CpG measurements were set to missing if detection p > 0.05 or <= 3 detection beads (M=66,532 probes). CpG sites were removed if >5% of samples were missing data. Based on the remaining CpG sites, samples were excluded if >5% of CpG sites had missing methylation values (N=43 removed). In the remaining samples, OOB background correction, RELIC dye bias correction, and RCP probe type bias correction were applied using Enmix.[3]

**Sample concordance**

Concordance between samples was measured using the SNP fingerprinting probes built into the array. Illumina SNP probes were converted to genotype data. These genotypes were assessed for pairwise IBD in PLINK. If a sample had a 100% match with an unexpected sample, the pair was compared against genotype array data. Under the assumption that array genotypes represented the 'truth' dataset, the sample from the pair that did not match the array genotype was excluded.

**Relatedness**

Kinship was determined from genotype data using KING.[4] For each pair of participants with estimated 3rd degree or closer relatedness, one was excluded, with the preference to retain cases. If case status matched, one of the pair was removed at random.
